# Incidence and epidemiology of Respiratory Syncytial Virus infections in children in rural Bangladesh: a prospective observational study

**DOI:** 10.1101/2025.07.22.25331975

**Authors:** Mohammad Shahidul Islam, Naito Kanon, Samin Huq, Mohammad Shameem Hassan, Shuborno Islam, Himadree Sarkar, Hafizur Rahman, Anannya Barman Jui, Sira Jam Munira, Yogesh Hooda, Justin Im, Andrea Haekyung Haselbeck, Deok Ryun Kim, Ju Yeon Park, Hyon Jin Jeon, Florian Marks, Samir K Saha, Senjuti Saha

## Abstract

**Background:** Respiratory syncytial virus (RSV) is a leading cause of acute respiratory infection (ARI) in young children worldwide, yet data from low- and middle-income countries (LMICs) are scarce. The data that are available are mostly hospital-based and urban. We conducted a prospective, community-based study to estimate the burden of RSV-associated ARI (RSV-ARI) among children <24 months in a rural area of Bangladesh.

**Methods:** The study was carried out in two villages within the Mirzapur Health and Demographic Surveillance System (HDSS) from August 2021 to June 2023. Village healthcare workers (VHWs) made weekly home visits to all registered households to identify ARI episodes in enrolled children <2 years, using the WHO ARI case definition. When ARI symptoms were reported, a nurse obtained written parental informed consent and collected a nasopharyngeal swab, which was tested for RSV by real-time RT-qPCR. We computed incidence per 1,000 child-years using Poisson regression, and estimated incidence rate ratios (IRRs) for male *versus* female and preterm *versus* term birth.

**Results:** 3,667 children contributed 3,008 child years of follow-up, 5,907 ARI episodes were recorded and 4,586 specimens collected and tested; a total of 7.1% (324/4,586) were RSV-positive. The overall RSV-ARI incidence was 107.7 per 1,000 child years (95% CI 96.6–120.1). In the first six months of life, the incidence was 164.5 per 1,000 child years, 1.5 times higher than the two-year average. Preterm infants had a 50% increased RSV-ARI risk (IRR 1.5; 95% CI 1.2–2.0). Cough was present in 95% of RSV cases, and chest indrawing, a sign not included in the WHO ARI definition, occurred in 9.8%.

**Conclusion:** RSV-ARI imposes a substantial community burden among rural Bangladeshi children under two years, especially in early infancy. These findings support the introduction of RSV-specific interventions (maternal and infant immunization or monoclonal antibodies) in similar rural LMIC settings.

## Introduction

Respiratory syncytial virus (RSV) is a leading cause of acute lower respiratory illness (ALRI) in children, especially during the first two years of life.^1,2^ Children in low- and middle-income countries (LMICs) carry a disproportionately high burden of severe RSV-associated illness.^3^ In 2019, an estimated 533,000 RSV-associated acute lower respiratory infection (RSV-ALRI) hospitalizations occurred globally, 93% of which were in LMICs.^1^ In the same year, countries with low socio-demographic index values recorded the highest age-standardized RSV mortality rate, estimated at 10.3 deaths per 100,000 population, compared to the global rate of 4.8 per 100,000. Among all regions, South Asia accounted for the highest number of RSV-related deaths.^4^

In Bangladesh, a country in South Asia, RSV poses a considerable health burden. A study in 2012 reported an incidence of 125 cases per 1,000 child-years among children under 24 months of age in Dhaka, the capital city.^5^ During the 2019 RSV season, about 20% of pediatric beds in a 650-bed tertiary-level hospital in Bangladesh were occupied by children with RSV infection, imposing substantial strain on hospital capacity and limiting the ability to manage other severe conditions.^6^ In a community-based study conducted by our group in three countries in South Asia, RSV was identified as the leading cause of neonatal infections in rural settings.^7^ However, little is known about its true community burden of RSV infection in Bangladesh’s rural areas, where laboratory and healthcare resources are limited.

High-income countries are beginning to deploy passive immunization (e.g., monoclonal antibodies and maternal vaccines) against RSV, but LMICs must set priorities against other public health needs. To guide evidence-based policy and optimize timing and targeting of interventions in Bangladesh, finely resolved, population-based burden data from rural communities are essential. This study therefore aims to quantify RSV associated acute respiratory infection (RSV-ARI) incidence, seasonality, and risk factors among children under two years in a rural Bangladeshi health demographic surveillance site.

## Methods

### Study Setting and Population

This prospective, community-based study was conducted over 23 months (August 2021– June 2023) in two unions, Bhatgram and Mirzapur Sadar, of the Mirzapur sub-district, Tangail District, Bangladesh. We enrolled permanent resident children aged <24 months through the Mirzapur Health and Demographic Surveillance System (MHDSS), a population-based platform that has been operational since 2007. Permanent residency was defined as the parents having resided in the study area for more than three months or for in-migrants, intending to remain for at least three months.

By 2022, the MHDSS covered ten unions (156 villages) with approximately 317,628 individuals under continuous surveillance. At that time, the crude birth rate was 17.3 per 1,000 population, the crude death rate was 7.0 per 1,000 population, and the under-five mortality rate was 30.3 per 1,000 live births. The two study unions together had 55,186 residents, yielding a population density of about 1,872 persons per square kilometer, and a birth rate of 15.5 per 1,000 people. Children <24 months residing in these unions were eligible for enrollment and followed until their second birthday, out-migration, deaths or study end, whichever occurred first.

### Case Identification and Sample Collection

Children under 24 months of age who were permanent residents of the study area were eligible for inclusion in the study. The World Health Organization (WHO) case definition for acute respiratory infection (ARI) was used for community-based surveillance of RSV infection. This definition includes acute illness, characterized by the sudden onset of symptoms, and respiratory infection, defined as the presence of shortness of breath, cough, sore throat or coryza.^8^ Local village health workers (VHWs) were trained to conduct weekly home visits and identify ARI based on the definition. Briefly, the study area was divided into 19 clusters, each consisting of approximately 250 households, with one VHW assigned per cluster. Each VHW visited 50 households per day, ensuring that all households in her area were visited once every week. VHWs asked the caregiver (primarily mothers) whether the child developed any sign of respiratory illness such as fever, cough, breathing difficulties, coryza or sore throat since the last visit to identify ARI cases in the community. All symptoms were recorded using a tablet-based data collection questionnaire. When an ARI case was identified through the system, the VHW triggered an automated electronic alert to our centralized coordination team. Nasopharyngeal swab samples were collected by trained nurses following standard procedures described below.

Within 72 hours of notification, a two-person sample collection team (one nurse, one porter) visited the household. After obtaining written informed consent from the child’s parent or guardian, the nurse collected a nasopharyngeal swab using a FLOQSwab (Copan Diagnostics, Italy). The swab was advanced into the nasopharynx, rotated 2–3 times in both directions, then placed in 1 mL Skim Milk–Tryptone–Glucose–Glycerol medium. Specimens were kept at 4–8 °C in a cool box and transported daily to the central laboratory for RSV testing. To avoid oversampling, children who had met the inclusion criteria and been swabbed were not eligible for repeat collection for 14 days after their initial sample.

### Sample processing

Nasopharyngeal specimens (140 µL) were processed for viral RNA extraction using the Quick-DNA/RNA Viral Magbead (Zymo Research, Orange, California) according to the manufacturer’s protocol. From each extraction, 7μl of extracted RNA underwent in RT-qPCR which was performed using the AgPath-ID^TM^ One-Step RT-qPCR kit (Thermo Fisher Scientific, Waltham, Massachusetts) and previously published RSV-specific primers and probe (Forward primer: - 5’-GGCAAATATGGAAACATACGTG AA-3’, reverse primer: 5’-TCTTTTTCTAGGACATTGTAYTGAACAG-3’, probe: 5’-CTGTGTATGTGGAGCCTTCGTGAAGCT-3’ (labeled at the 5’ end with FAM and quenched at the 3’end with Black Hole Quencher-1)). Reactions were run on an Applied Biosystem qPCR 7500 Fast Dx system (Thermo Fisher Scientific, Waltham, MA, USA). Cycling conditions were 50°C for 10 minutes, 95°C for 1 minute, and 40 cycles of 95°C for 10 seconds and 60°C for 1 minute. Specimens were considered RSV-positive if they produced a true sigmoidal amplification curve with a cycle threshold (Ct) value <35.

### Statistical analysis

We calculated RSV-ARI incidence using Poisson regression, with each child’s follow-up time as the exposure. Child-specific follow-up began at enrollment and ended at age 24 months, out-migration, death, or study end, whichever came first. Person-time was computed as the difference between each child’s first and last home-visit dates (or date of death, where applicable), expressed in child-years. Unique ARI episodes were defined as those separated by >14 days. We estimates incidence per 1,000 child-years, along with 95% confidence intervals (CIs). We also estimated incidence rate ratios (IRRs) and 95% CIs for key risk factors: age group (0–5, 6–11, 12––23 months), sex, and preterm birth status (<37 weeks’ gestation, ascertained from last menstrual period data collected every 16 weeks in the HDSS).

Socioeconomic status (SES) was derived via principal component analysis of household asset variables; households were ranked by their first-component scores and categorized into quintiles (from lowest to highest). We assessed the association between SES quintile and RSV-ARI incidence by including quintile as a categorical predictor in the Poisson models.

All analyses were conducted in Stata version 14.0 (StataCorp, College Station, TX) and R version 4.2.2 (R Foundation for Statistical Computing, Vienna, Austria). Statistical significance was set at a two-sided *p*-value <0.05.

## Ethical considerations

We obtained ethical clearance for this study from the Ethical Review Board of Bangladesh Shishu Hospital and Institute, Bangladesh and from the Institutional Review Board of the International Vaccine Institute (IVI IRB), South Korea. Written informed consent was obtained from caregivers for participation in the study. Additionally, the sample collection team obtained separate written consent for the collection of nasopharyngeal samples. Participation in this study was entirely voluntary, and no incentives were provided to caregivers. However, children enrolled in the study received free medical treatment for any illness through the outpatient department of Kumudini Women’s Medical College Hospital.

## Results

Between August 2021 and June 2023, we approached 3,668 children under 24 months of age for enrollment, and 3,667 were successfully enrolled (Figure 1). One child died before the VHW could reach the household. Over the 23 month period, VHWs completed 143,918 weekly household visits; enrolled children were present at 136,935 (95.2%) of these visits. During these visits, 6,992 episodes of illness were recorded, of which 5,907 episodes met the WHO ARI case definition.

**Figure 1:**
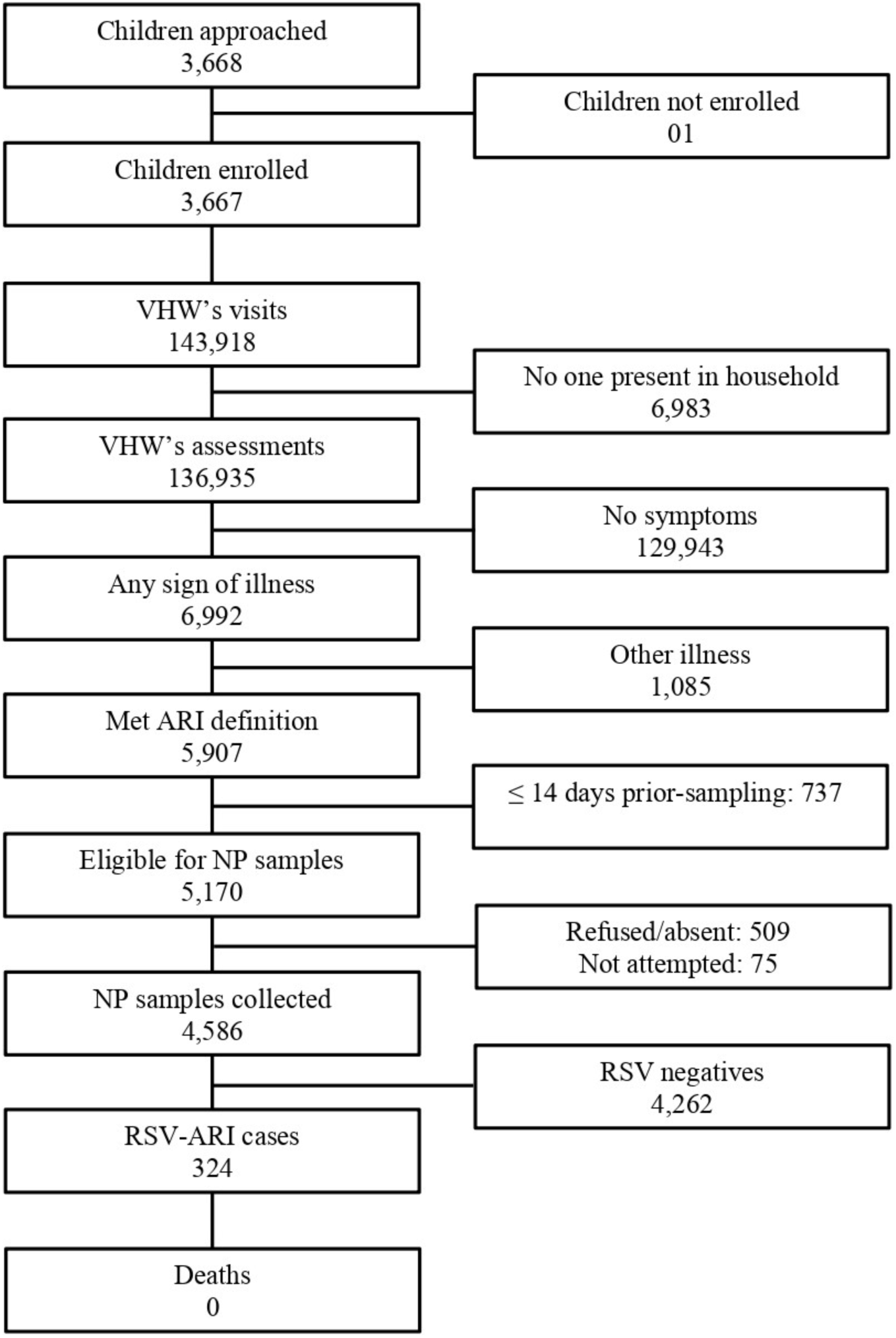
Study flow diagram of the community-based RSV-ARI surveillance in Mirzapur, Bangladesh

Nasopharyngeal swabs were obtained for RSV testing for 77.6% (4,586/5,907) of the ARI episodes. We excluded 737 ARI episodes from sampling as samples had been collected within the prior 14 days. Additionally, in 509 ARI episodes, samples could not be collected because the child was absent, or the caregiver refused to provide consent. In 75 episodes, the sample collection team could not visit the household in time due to vairous logistical challenges. RSV was detected by RT-qPCR in 324 samples (7.1%, 324/4,586), representing 316 children, eight of whom had two distinct RSV infections. No RSV-associated deaths occurred during the course of this study.

The demographic and socio-economic characteristics of enrolled children are summarized in Table 1. Of the 3,667 enrolled children, 1,875 (51.1%) were male and 1,792 (48.9%) were female. Age at enrollment was skewed toward younger infants: 30.1% (1,104/3,667) were enrolled within their first two months of life, 49.0% (1,798/3,667) by five months, and 70.6% (2,589/3,667) by 12 months; the remaining 29.4% (1,078/3,667) were enrolled between 12 and 24 months. Preterm births (<37 weeks’ gestation) accounted for 17.3% (483/2,785) of the cohort; 82.7% (2,302/2,785) were term. Mothers’ median age was 25 years (IQR: 20–30); 84.4% (2,909/3,446) were homemakers, 49.6% (1,709/3,446) had only primary education, and 41.6% (1,435/3,446) had secondary or higher education.

**Table 1:** Characteristics of the study participants by RSV infection status.

| Characteristics | Overall | % | RSV+ | % | RSV- | % | p-value |
| --- | --- | --- | --- | --- | --- | --- | --- |
| Children | 3,667 |  | 316 |  | 3,351 |  |  |
| Years contributed to the study (years) | 3,008 |  | 352 |  | 2,656 |  |  |
| Gestational Age (n=2785 available) | 2,785 |  | 276 |  | 2509 |  | 0.000 |
| Preterm (32-36) | 483 | 17.3 | 69 | 25.0 | 414 | 16.5 |  |
| Term (37+) | 2,302 | 82.7 | 207 | 75.0 | 2,095 | 83.5 |  |
| Age at enrollment <28 days | 637 | 17.4 | 71 | 22.5 | 566 | 16.9 | 0.012 |
| Age at enrollment >=28 to <59 days | 467 | 12.7 | 54 | 17.1 | 413 | 12.3 | 0.015 |
| Age at enrollment >=59 days to <5 months | 694 | 18.9 | 75 | 23.7 | 619 | 18.5 | 0.022 |
| Age at enrollment >=5 months to <12 months | 791 | 21.6 | 79 | 25.0 | 712 | 21.2 | 0.121 |
| Age at enrollment >12 months to <24 months | 1,078 | 29.4 | 37 | 11.8 | 1041 | 31.1 | 0.000 |
| Male Sex | 1,875 | 51.1 | 172 | 54.4 | 1,703 | 50.8 | 0.219 |
| Female | 1,792 | 48.9 | 144 | 45.6 | 1,648 | 49.2 |  |
| <u>Maternal</u> |  |  |  |  |  |  |  |
| Median Mother's age (IQR) at childbirth (n=3446) | 25 [20-30] |  | 25 [20-30] |  | 25 [20-30] |  | 0.799 |
| Homemaker (n=3446) | 2,909 | 84.4 | 266 | 86.4 | 2,643 | 84.2 | 0.324 |
| Completion of Primary Education (n=3446) | 3,114 | 91.2 | 277 | 90.0 | 2,867 | 91.4 | 0.397 |
| Delivery place (n=2785) | 2,785 |  | 276 |  | 2,509 |  | 0.671 |
| Facility Delivery | 2,471 | 88.7 | 247 | 89.5 | 2,224 | 88.6 |  |
| Home Delivery | 314 | 11.3 | 29 | 10.5 | 285 | 11.4 |  |
| <u>Demographic</u> |  |  |  |  |  |  |  |
| Socioeconomic Status (n=3106) | 3,106 |  | 278 |  | 2,828 |  |  |
| Lowest | 635 | 20.4 | 63 | 22.7 | 572 | 20.2 | 0.337 |
| Lower | 618 | 19.9 | 65 | 23.4 | 553 | 19.6 | 0.127 |
| Middle | 614 | 19.8 | 47 | 16.9 | 567 | 20.0 | 0.209 |
| Higher | 622 | 20.0 | 60 | 21.6 | 562 | 19.9 | 0.497 |
| Highest | 617 | 19.9 | 43 | 15.5 | 574 | 20.3 | 0.054 |
| Sources of Drinking (n=3128) | 3,128 |  | 281 |  | 2,847 |  |  |
| Piped Water | 9 | 0.3 | 0 | 0.0 | 9 | 0.3 | 0.345 |
| Well-Water | 3,001 | 95.9 | 275 | 97.9 | 2,726 | 95.7 | 0.087 |
| Surface Water | 1 | 0.03 | 1 | 0.4 | 0 | 0.0 | 0.002 |
| Filtered Water | 115 | 3.7 | 5 | 1.8 | 110 | 3.9 | 0.077 |
| Other | 2 | 0.1 | 0 | 0.0 | 2 | 0.1 | 0.657 |
| Received at least one vaccine from epi program (n=3502) | 3,436 | 98.1 | 307 | 99.4 | 3,129 | 98.0 | 0.094 |
| Illness history |  |  |  |  |  |  |  |
| Number of household visit | 136,935 |  | 16,357 |  | 120,578 |  |  |
| Number of visits per child | 55 [37-68] |  | 63 [47-74] |  | 53 [36-67] |  | 0.000 |
| Number of visits with any illness sign | 6,992 | 5.1 | 1636 | 10.0 | 5,356 | 4.4 | 0.000 |
| Number of visits with ARI illness sign | 5,907 | 84.5 | 1482 | 90.6 | 4,425 | 82.6 | 0.000 |
| Number of ARI per child | 4 [2-8] |  | 7 [4-12] |  | 3 [2-6] |  | 0.000 |

The enrolled children contributed a total of 3,008 child-years, resulting in an overall incidence of 107.7 RSV-ARI per 1,000 child-years (95% Confidence Interval (CI): 96.6-120.1). Incidence was highest in the first six months of life at 164.5 per 1,000 child-years (95% CI: 134.2-201.6), declined to 150.1 (95% CI:126.2-180.0) between 6-11 months, and further to 67.1 (95% CI:55.6-80.8) at 12-23 months (Figure 2).

**Figure 2:**
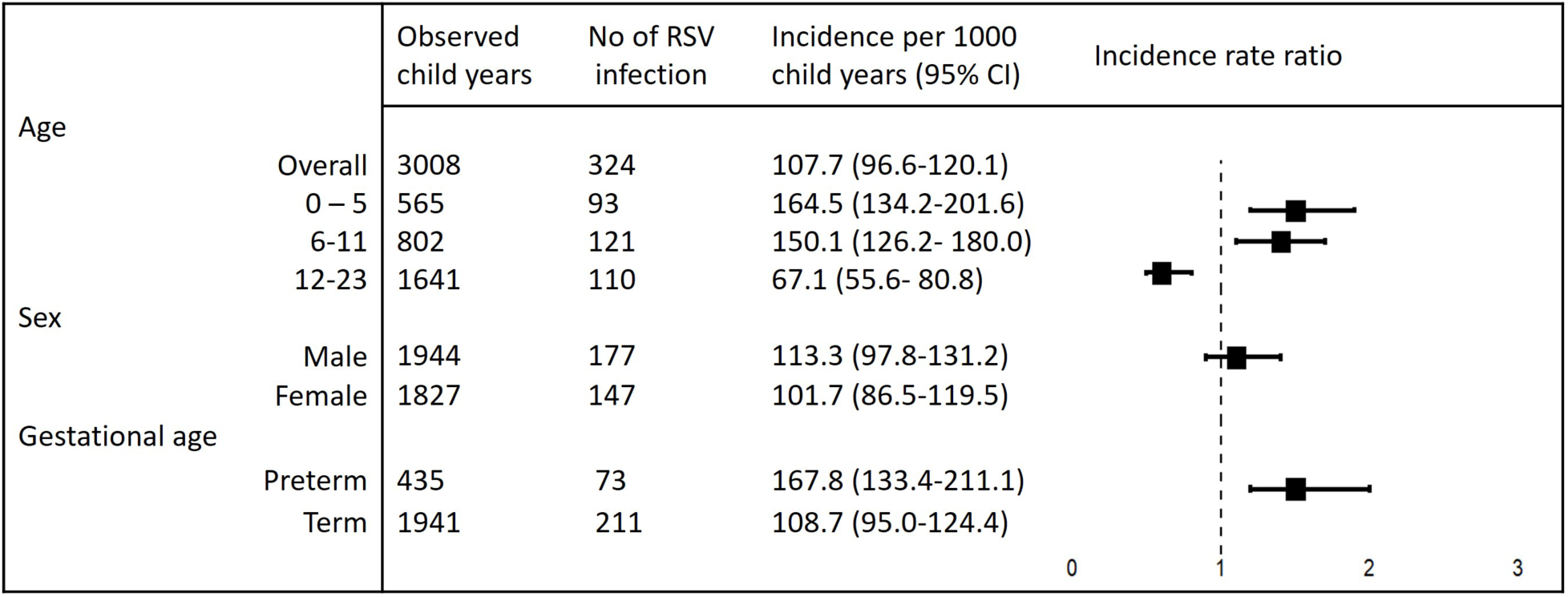
Incidence and rate ration of RSV-ARI in rural Bangladesh by child age, sex and gestational age

Preterm infants experienced an RSV-ARI incidence of 167.8 per 1,000 child-years (95% CI: 133.4–211.1), which was 1.5 times higher than the incidence among term infants, 108.7 per 1,000 child-years (95% CI: 95.0–124.4). There was no significant difference in RSV-ARI incidence by sex; males had an RSV-ARI incidence of 113.3 per 1,000 child-years (95% CI: 97.8-131.2), compared to 101.7 per 1,000 child-years in females (95% CI: 86.5-119.5) (Figure 2).

RSV-ARI incidence exhibited clear seasonal peaks (Figure 3). In August–September 2021, 37.6% (92/245) of swabs were RSV-positive; a second peak spanned October 2022 to February 2023, with 16.1% (196/1,218) positivity. During the remaining months, RSV detection fell to near zero.

**Figure 3:**
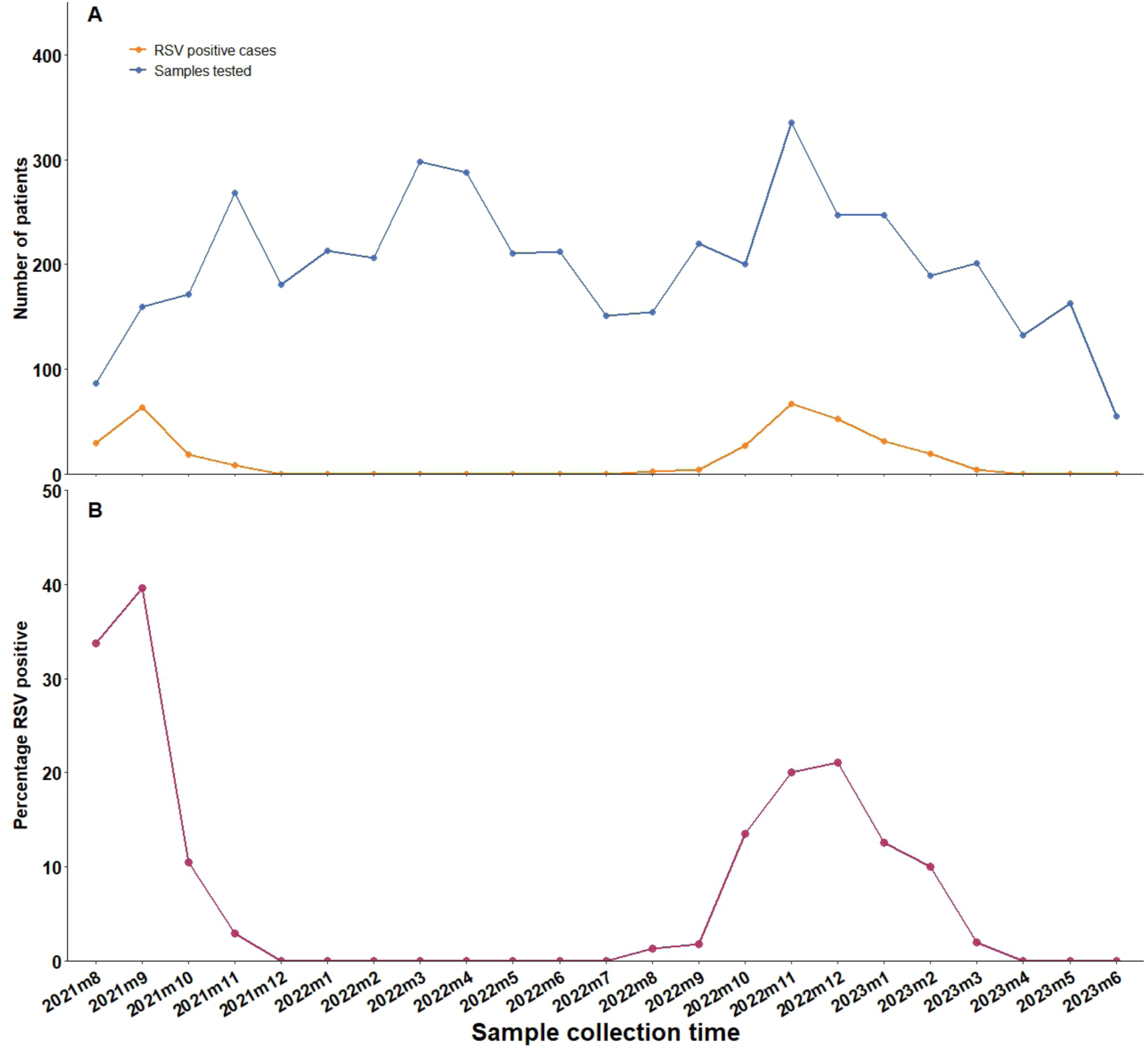
Monthly RSV detection rate in nasopharyngeal samples from children in rural Bangladesh. (A) Number of nasopharyngal samples tested and RSV cases detected in each calendar months, (B) Parcentage of RSV positive cases in nasopharyngeal samples in each calendar months

Among the 324 RSV-positive episodes, cough was nearly universal (95.7%, 310/324), followed by breathing difficulties (32.7%, 106/324)), fever (26.2%, 85/324), and fast breathing (26.1%, 83/324) (Table 2). Chest indrawing, a marker of more severe lower respiratory involvement not included in the WHO ARI definition, occurred in 9.8% (31/324) of RSV cases versus 2.3% (91/4,262) of RSV-negative ARI episodes.

**Table 2:** Clinical characteristics of children with RSV-ARI in rural Bangladesh.

| Cinical Sign | Overall (%)<br>n=4,586 | RSV Positive (%)<br>n=324 | RSV Negative (%)<br>n=4,262 | p-value |
| --- | --- | --- | --- | --- |
| Cough | 3,889 (84.8) | 310 (95.7) | 3,579 (83.9) | 0.000 |
| Chest Indrawing | 122 (2.9) | 31 (9.8) | 91 (2.3) | 0.000 |
| Difficulties in Breathing | 956 (20.9) | 106 (32.7) | 850 (19.9) | 0.000 |
| Fast Breathing | 485 (11.4) | 83 (26.1) | 402 (10.3) | 0.000 |
| Fever | 731 (15.9) | 85 (26.2) | 646 (15.2) | 0.000 |
| Sore Throat | 22 (0.5) | 3 (0.9) | 19 (0.5) | 0.228 |

*\*\*Chest Indrawing and Fast Breathing was available for 4,236 cases*

## Discussion

In this prospective, 23-month, community-based study conducted in rural Bangladesh, we followed 3,667 children to estimate the burden of RSV-ARI. The study was conducted in the context of the recent licensure of a maternal RSV vaccine and monoclonal, which holds promise for reducing RSV-associated illness at the community level. ^9–11^ Overall, 7.1% of ARI cases tested positive for RSV, corresponding to an overall RSV-ARI incidence rate of 107.7 episodes per 1,000 child-years. Two distinct seasonal peaks in RSV activity, August–September 2021 and October 2022–February 2023, accounted for virtually all RSV detections.

Our overall RSV-ARI incidence aligns closely with previous findings from urban populations in Bangladesh. A study among children under the age of 2 years in urban Dhaka reported an RSV incidence of 125 episodes per 1,000 child-years.^5^ Another study reported an incidence of 112.4 cases in urban Dhaka of Bangladesh.^12^ Similarly, we and others have documented substantial RSV hospital burden in Bangladesh’s hospitals.^6,13^ Compared with other South Asian settings, our incidence is lower than the 213 per 1,000 child-years reported among 0–6-month-olds in rural Nepal.^14^ This difference may be attributed to the highly seasonal nature of RSV,^15^ as well as variation in observation periods and study populations.^16^ The Nepal study followed children only through the first six months of life, a period when susceptibility to RSV is highest.^17,18^ In our cohort, we also observed the highest incidence of RSV-ARI during the first six months of life (164.5 episodes per 1,000 child-years). Conversely, our rates exceed those from a remote tribal Indian population, where infants 0–3 months experienced 12.7 RSV LRTI episodes per 1,000 child-years.^19^ Heterogeneity in study design, surveillance intensity, and case definitions must be considered when comparing across regions.

Taken together, these findings suggest that RSV prevention strategies including maternal immunization and immunoprophylaxis using monoclonal antibodies for high-risk populations are likely to have comparable effectiveness across both rural and urban settings in the country. Notably, RSV-ARI incidence peaked at 164.5 per 1,000 child-years in the first six months of life,^7^ a window ideally covered by maternal antibody transfer.^9,11,20^ In our cohort, 17.3% of children were born preterm, similar to the national rate of 16.2%.^21^ Preterm infants faced a 1.5-fold higher RSV-ARI risk than term peers (IRR 1.5; 95% CI: 1.2–2.0),

The markedly elevated risk in preterm infants is consistent with global literature,^2^ including studies from South Asia.^14^ Preterm children are at increased risk of severe RSV illness due to reduced transplacental transfer of maternal antibodies and immature lung development.^20^ Therefore, optimizing the timing of maternal RSV vaccination is required to protect preterm children the most vulnerable group to RSV infection. Vaccinating mothers earlier in pregnancy may provide better protection against RSV for preterm children, who are the most susceptible to RSV infection. However, this strategy needs careful consideration, as earlier immunization may result in waning immunity by the time a term children is born, potentially necessitating a booster dose to maintain protection.^22^ Alternatively, passive immunoprophylaxis with monoclonal antibodies (e.g. nirsemivab) should be considered a priority intervention for this high-risk group to ensure robust protection across the full birth spectrum.^23,24^ Furthermore, modeling studies have projected that newer long-acting monoclonal antibodies such as nirsevimab could confer broad, season-long protection for all infants, potentially yielding a much broader impact on RSV burden in our population than maternal vaccination alone.^6^

Consistent with all previous studies that have reported seasonal variation in RSV infection burden,^17^ our data show that RSV incidence varied by month. During our study period, with the highest burden observed during the fall season (September to November). Unlike countries in temperate regions, with well defined RSV seasons, equatorial countries such as Bangladesh experience varied RSV peaks, sometimes in winter, summer, or even twice a year. ^25,26^

Although hospital-based studies have suggested higher RSV severity and care-seeking among male children, we observed no significant difference in community-based RSV-ARI incidence between sexes (male: 113.3 vs. female: 101.7 per 1,000 child-years).^27^ In Bangladesh, in-hospital data show sex-based disparities in pneumonia severity and mortality, despite equivalent treatment access, largely driven by differential care-seeking behaviors.^28^ Our population-based surveillance, by contrast, indicates that male and female infants are equally susceptible to RSV infection, reinforcing the need for prevention strategies that reach all children irrespective of sex.

Although RSV symptoms overlap with those of other viral infections,^29^ our study identified chest indrawing as a prominent clinical sign associated with RSV infection. Other previous reports also suggested that chest indrawing was consistently associated with RSV infection.^30^ This symptom is not currently included in the WHO case definition for RSV-ARI.^8^ Given its strong association with RSV and its feasibility for detection by trained community health workers,^31^ we recommend its inclusion in community-based RSV surveillance protocols.

Our study offers several distinctive strengths. First, by embedding surveillance within a rural HDSS where diagnostic capacity is limited and RSV testing is not routine, we provide the first population-level estimate of RSV burden in this setting, complementing prior work that followed a small cohort of 252 children and reported only pneumonia cases.^32^ Second, our weekly household visits by trained village health workers, coupled with a mobile specimen-collection team of nurses and porters, minimized barriers to care and ensured timely capture of symptomatic ARI and high-quality respiratory sampling directly in the home. Third, we enrolled every resident child under two years regardless of their age at enrollment, thereby capturing all eligible RSV-ARI episodes across the entire study area over 23 months.

Nevertheless, our findings should be interpreted in light of several limitations. We were unable to collect specimens for approximately 10% of identified ARI episodes, mainly due to caregiver refusal or child absence, which may have led to some underestimation of RSV incidence. Weekly surveillance, while intensive, may have missed very brief illnesses occurring between visits. Finally, the longstanding presence and health-education activities of our VHWs could have influenced caregiver behavior and care-seeking, potentially altering the natural history and reporting of RSV infections in these communities.

In summary, our community-based surveillance confirms a substantial burden of RSV-ARI in rural Bangladesh, with incidence peaking in the first six months of life and disproportionately affecting preterm infants. Chest indrawing, a sign not currently in the WHO ARI definition, was observed in nearly 10% of RSV cases and may improve case detection if added to surveillance protocols. The most critical priority is implementation of prevention strategies. We recommend that policymakers implement maternal RSV vaccination to protect all newborns and deploy monoclonal antibody prophylaxis for the highestrisk infants, ensuring comprehensive protection across the full birth spectrum.

## Data Availability

All data produced in the present study are available upon reasonable request to the authors.

## Contributors

MSI was responsible for data validation and analysis of the study, and drafting the initial manuscript. NK contributed to data management, analysis and drafting the manuscript. SH contributed to study coordination, training and data analysis. MSH was involved in coordinating the field implementation of the study and sample collection. SI HR and HS involved in sample processing and testing. ABJ and SJM was involved in coordination and trained doctors and nurses, assisted in the implementation of the study, and contributed to manuscript editing. YH participated in data analysis, preparation of figures, and manuscript editing. SKS and SS contributed to conceptualization, funding acquisition, study design, implementation, monitoring, data analysis, drafting the initial manuscript, and preparation of figures. FM, JI, JYP, AH, DRK contributed to conceptualization, funding acquisition, study design and monitoring, of the project. All authors reviewed the manuscript and agreed on the contents.

## Conflict of interest

The authors have declared no conflict of interest.

### Declaration of Generative AI and AI-assisted technologies in the writing process

During the preparation of this work the authors used ChatGPT to check grammar, spellings, and improve readability of the manuscript. After using this tool/service, the authors reviewed and edited the content as needed and take full responsibility for the content of the publication.

